# Leveraging molecular-QTL co-association to predict novel disease-associated genetic loci using a graph convolutional neural network

**DOI:** 10.1101/2024.03.04.24303678

**Authors:** Julian Ng-Kee-Kwong, Andrew D. Bretherick

**Author notes:** Corresponding author(s). E-mail(s); Contributing authors. These authors contributed equally to this work.

## Abstract

Genome-wide association studies (GWAS) have successfully uncovered numerous associations between genetic variants and disease traits to date. Yet, identifying significantly associated loci remains a considerable challenge due to the concomitant multiple-testing burden of performing such analyses genome-wide. Here, we leverage the genetic associations of molecular traits – DNA CpG-site methylation status and RNA expression – to mitigate this problem. We encode their co-association across the genome using PinSage, a graph convolutional neural network-based recommender system previously deployed at Pinterest. We demonstrate, using this framework, that a model trained only on methylation quantitative trait locus (QTL) data could recapitulate over half (554,209/1,021,052) of possible SNP-RNA associations identified in a large expression QTL meta-analysis. Taking advantage of a recent ‘saturated’ map of height associations, we then show that height-associated loci predicted by a model trained on molecular-QTL data replicated comparably, following Bonferroni correction, to those that were genome-wide significant in UK Biobank (88% compared to 91%). On a set of 64 disease outcomes in UK Biobank, the same model identified 143 independent novel disease associations, with at least one additional association for 64% (41/64) of the disease outcomes examined. Excluding associations involving the MHC region, we achieve a total uplift of over 8% (128/1,548). We successfully replicated 38% (39/103) of the novel disease associations in an independent sample, with suggestive evidence for six additional associations from GWAS Catalog. Replicated associations included for instance that between rs10774625 (nearest gene: SH2B3/ATXN2) and coeliac disease, and that between rs12350420 (nearest gene: MVB12B) and glaucoma. For many GWAS, attaining such an enhancement by simply increasing sample size may be prohibitively expensive, or impossible depending on disease prevalence.

## Introduction

Genome-wide association studies (GWAS), which are designed to identify trait-associated genetic variation, have been performed for many different traits: both non-molecular (e.g. height, cardiovascular disease) and molecular traits (e.g. DNA CpG-site methylation status, RNA expression). However, the statistical power of many disease GWAS is limited by disease prevalence, and by the logistical and financial demands associated with recruiting large cohorts.

In recent years, graph convolutional neural networks have been used in many applications, including as part of web-scale ‘recommender systems’ [1, 2]. Recommender systems are tools designed to make suggestions that are of greatest interest to a given user. A typical application may be to guide a user, based on their purchasing history, to other items they might like. Such recommender systems may function by generating an embedding – a multi-dimensional representation – of the items based on the activity of large sets of users. This embedding encapsulates existing knowledge about a domain, capturing for instance the similarity between items based on user interaction. Here, we apply this framework to the genetic associations of molecular traits and use it to identify novel disease associations based on the loci already known to be associated with a given disease.

We hypothesised that co-association of two genetic loci with a molecular trait was likely to be informative with respect to their co-association with other traits. Using the Deep Graph Library (DGL) implementation [3] of PinSage [4], a recommender system that has previously been deployed at Pinterest (https://www.pinterest.co.uk/), we encode the knowledge contained in molecular trait (DNA CpG-site methylation status and RNA expression) GWAS and use it to augment existing disease GWAS results.

We first demonstrate that our embeddings are biologically meaningful in three stages: 1) We show that our model can successfully predict co-association between SNP and DNA CpG-site methylation status (henceforth referred to as SNP-CpG associations); 2) We recapitulate, using a model trained only on methylation QTL (meQTL) data, the associations between SNP and RNA expression (henceforth referred to as SNP-RNA associations) identified in a large expression QTL (eQTL) meta-analysis of 31,684 individuals [5]; and 3) We interrogate the molecular-QTL network, showing that SNP or node ‘importance’, as measured by eigenvector centrality, is associated with an increased likelihood of disease association.

Having demonstrated the above, we then use these embeddings to predict novel height-associated genetic loci. We show that 88% (23/26) of the independent novel loci thus identified, that were present in a ‘saturated’ map of height associations [6], successfully replicate; this is compared to 91% (897/984) of those that were genome-wide significant (p-value < 5 × 10^−8^) in UK Biobank, accounting for multiple-testing via Bonferroni correction in both cases. Finally, we apply our approach to a set of self-reported disease outcomes in UK Biobank. We identify at least one additional disease-associated locus in almost two-thirds of disease traits analysed (41/64). Overall, our model achieves an uplift of about 8% compared to the number of genome-wide significant associations considered (128/1,548) – excluding here those involving the MHC region. We successfully replicate 38% (39/103) of those associations that were testable in FinnGen [7], and find suggestive evidence for six additional disease associations in GWAS Catalog. Given the potential to enhance discovery of disease-associated genetic loci, and the benefits of genetic evidence in supporting drug targets [8], we propose the future use of such recommender systems as a standard part of post-GWAS analysis.

## Results

### Biologically Meaningful Embeddings

Before presenting the results when predicting held-out SNP-CpG associations and SNP-RNA associations (Model 1 and Model 2, respectively; See Methods), we first outline how recommendations are made and how model performance is assessed.

In brief, we make recommendations based on a ‘query’ SNP – here, we select as the query SNP that which is most significantly associated (smallest p-value) with the molecular trait (DNA CpG-site methylation status or RNA expression). Once selected, its similarity to all other SNPs is computed and the resulting list ordered. A set of ‘K’ recommendations is then made by taking the first ‘K’ elements of the ordered list, which are effectively the K-nearest neighbours of the query SNP in the embedding space. The process of making recommendations is more fully described in the Methods. For a given trait (e.g. methylation status of a specific CpG-site), we define a set of recommendations as a ‘hit’ if it contains at least one SNP that is associated with the trait at a genome-wide significant threshold (p-value < 5 × 10^−8^). For a set of traits (e.g. the set of all queried CpG-sites in Model 1), the ‘hit-rate’ is then the fraction that are deemed ‘hits’.

#### Model 1: Predicting SNP-CpG associations

We demonstrate that the genetic variant most significantly associated with the methylation status of a given DNA CpG-site can be used to predict, for that CpG-site, other SNP associations that were held-out during model training (the ‘test set’): Table 1. We consider here only those CpG-sites having at least one SNP association in the test set. When making 10 recommendations based on the most significantly associated SNP, our model achieves a hit-rate of 54%, which goes up to 90% when the number of recommendations is increased to 100. Having obtained these results in the standard embedding (r^2^ < 0.9 between SNPs; minor allele frequency (MAF)> 0.05), we proceeded to demonstrate comparable hit-rates in an embedding limited to a set of pseudo-independent SNPs (r^2^ < 0.05; MAF> 0.01), indicating that the results observed are not simply a trivial recapitulation of the linkage disequilibrium (LD) structure surrounding the lead SNP. Model performance over time is shown in Supplementary Figure S1.

**Table 1.** Results when predicting meQTL test data. 1, 10, 100: the total number of hits (hit-rate) when 1, 10 or 100 recommendations are made. N*_CpG_*: the number of CpG-sites present in the test set. n*_assoc_*: the median (IQR) number of SNP associations, per CpG-site, present in the test set. N*_assoc_*: the total number of SNP-CpG associations present in the test set. Model trained on 81% of the available meQTL data. Test data not seen during training. Standard embedding: r^2^ < 0.9, MAF> 0.05; Pseudo-independent embedding: r^2^ < 0.05, MAF> 0.01. r^2^ and MAF: LD threshold and MAF thresholds of the SNPs present in the embedding. 952,129 and 91,722 SNPs are present in the standard and pseudo-independent embeddings, respectively.

| Embedding | 1 | 10 | 100 | $N_{CpG}$ | $n_{assoc}$ | $N_{assoc}$ |
| --- | --- | --- | --- | --- | --- | --- |
| Standard | 21,762 (16%) | 73,558 (54%) | 122,708 (90%) | 137,030 | 4 (2-8) | 998,379 |
| Pseudo-independent | 6,974 (22%) | 22,586 (71%) | 27,866 (88%) | 31,677 | 1 (1-2) | 42,818 |

#### Model 2: Predicting SNP-RNA associations

We show that the co-association information of DNA CpG-site methylation status can be used to effectively predict SNP-RNA associations. We perform a similar analysis to that above, but this time training on all of the meQTL data, and testing on eQTL data. We also include an ‘adjusted hit-rate’ whereby we make the same number of recommendations per RNA as there are SNPs associated to the RNA in the test set (Table 2). We show, based on this ‘adjusted hit-rate’, that we successfully recapitulate, excluding the lead SNP used to query the embedding, over half (54%; 554,209/1,021,052) of all possible SNP-RNA associations present in eQTLGen [5], without using any eQTL data during model training. We again demonstrate, using our pseudo-independent embedding, that our results are not simply explained by the LD structure surrounding the query SNP.

**Table 2.** Results when predicting eQTL data. 1, 10, 100: the total number of hits (hit-rate) when 1, 10 or 100 recommendations are made. ‘Adjusted’: the total number of hits (hit-rate) when the number of recommendations made for a given gene was set to the number of SNP associations for that gene. N*_genes_*: the number of genes present in the test set. Model trained on 100% of the available meQTL data. n*_assoc_*: the median (IQR) number of SNP associations, per RNA, present in the test set. N*_assoc_*: the number of SNP-RNA associations present in the test set. Standard embedding: r^2^ <0.9, MAF> 0.05; Pseudo-independent embedding: r^2^ < 0.05, MAF> 0.01. r^2^ and MAF: LD threshold and MAF thresholds of the SNPs present in the embedding.

| Embedding | 1 | 10 | 100 | ‘Adjusted’ | $N_{genes}$ | $n_{assoc}$ | $N_{assoc}$ |
| --- | --- | --- | --- | --- | --- | --- | --- |
| Standard | 8,242 (55%) | 13,070 (88%) | 14,687 (98%) | 554,209 (54%) | 14,935 | 46 (17-94) | 1,021,052 |
| Pseudo-independent | 4,287 (43%) | 8,900 (90%) | 9,734 (98%) | 25,723 (48%) | 9,926 | 4 (2-7) | 53,962 |

Finally, we attempted to predict, using the standard embedding, *trans* associations (defined as eQTLs located > 5Mb distal to the gene) based on the lead *cis*-eQTL (defined as < 1Mb from the gene). When the number of recommendations per RNA was again set to the number of SNPs associated in *trans* to the RNA in the test set, our model performed significantly better than chance (29 against 17, out of 6,690 possible *trans* associations for 1,965 genes; p-value = 0.0044), although this improvement in performance was likely constrained by the sparsity of *trans* associations reported in eQTLGen (only around 2,600 *trans* associations tested per RNA, after limiting eQTLs to those SNPs present in our embedding). Of note to the reader, we use the terms ‘RNA’ and ‘gene’ interchangeably throughout this manuscript.

#### Model 3: Node importance linked to disease association

The importance of a node within a network can be measured in many ways. Here, we measure node importance using eigenvector centrality after computing the similarity matrix based on the pseudo-independent embedding. We compare a set of disease-associated SNPs (n = 319) to a set of MAF-matched control SNPs; the difference, assessed here using the Mann-Whitney U test, was strongly significant (p-value = 4.2 × 10^−12^; Figure 1). When disease categories were considered separately, differences remained significant for autoimmune diseases, diabetes, obesity and ‘other’ diseases following Bonferroni correction per disease category (Supplementary Table 1). For the remaining categories, the disease group was still associated with higher eigenvector centrality, though not statistically significant. Overall, this suggests an informative relationship between centrality within the molecular-QTL network and disease association.

**Fig. 1.**
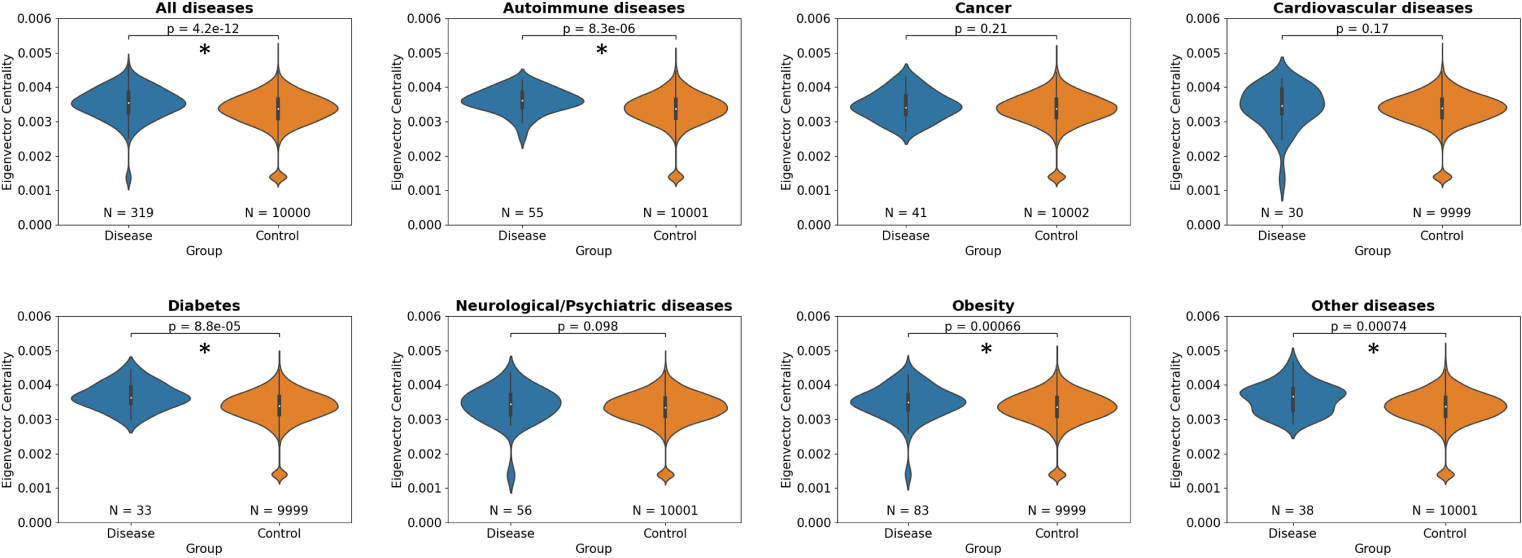
Centrality within the molecular-QTL network is linked to likelihood of disease association. Node importance, as measured by eigenvector centrality, was compared between SNPs associated with disease and a set of MAF-matched control SNPs (Methods). Uncorrected p-values from Mann-Whitney U test are shown. The number of SNPs considered is provided below each violin plot. Differences that remained significant following Bonferroni correction are indicated by an asterisk.

### Augmentation of Existing GWAS

Having demonstrated that the models are biologically meaningful, we applied a similar approach to height and a set of 64 self-reported disease outcomes in UK Biobank. We hypothesised that the models had learnt a genome-wide map of the functional interactions between SNPs (Figure 2), which we leverage here in order to identify additional SNP associations for each of the traits considered. Full detail of how the recommendations were made for each GWAS is provided in the Methods, and a summary is presented in Algorithm 1. By design, none of the novel loci presented were genome-wide significant in the original GWAS in UK Biobank, neither were they in linkage disequilibrium with any of them (r^2^ < 0.05).

**Fig. 2.**
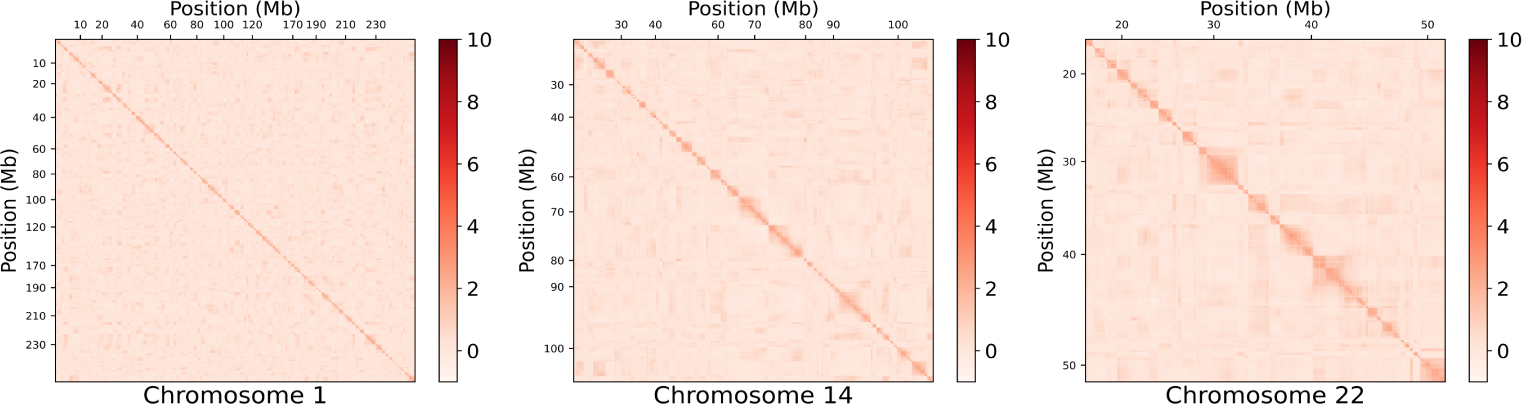
Heatmap of the similarity matrix representing functional interaction across SNPs. Presented here are heatmaps for three representative chromosomes: 1, 14 and 22. Genome position is on both axes. Higher values (darker red) indicate greater similarity between two loci, similarity here being given by the dot product of their respective embeddings. The diagonal represents self-similarity. High similarity scores closer to the diagonal represent the local (*cis*) region; whilst those further away from the diagonal represent *trans*-region(s) co-associating together. The similarity matrix is based on the pseudo-independent embedding. Heatmaps for the remaining chromosomes are provided in Supplementary Figure S2.

We first demonstrated that the augmented GWAS results of height in UK Biobank replicate as expected in a very large meta-analysis where the contribution of UK Biobank was removed (See Methods) [6]. After identifying 107 independent novel height-associated loci, we attempted replication of 26 such loci for which the SNP was included in the replication cohort. 88% (23/26) replicated successfully, with a consistent direction-of-effect (Supplementary Table 2). In comparison, 91% (897/984) of those that were genome-wide significant in UK Biobank replicated in the meta-analysis. Multiple-testing was accounted for via Bonferroni correction in both cases.

#### Algorithm 1 Identifying novel disease associations

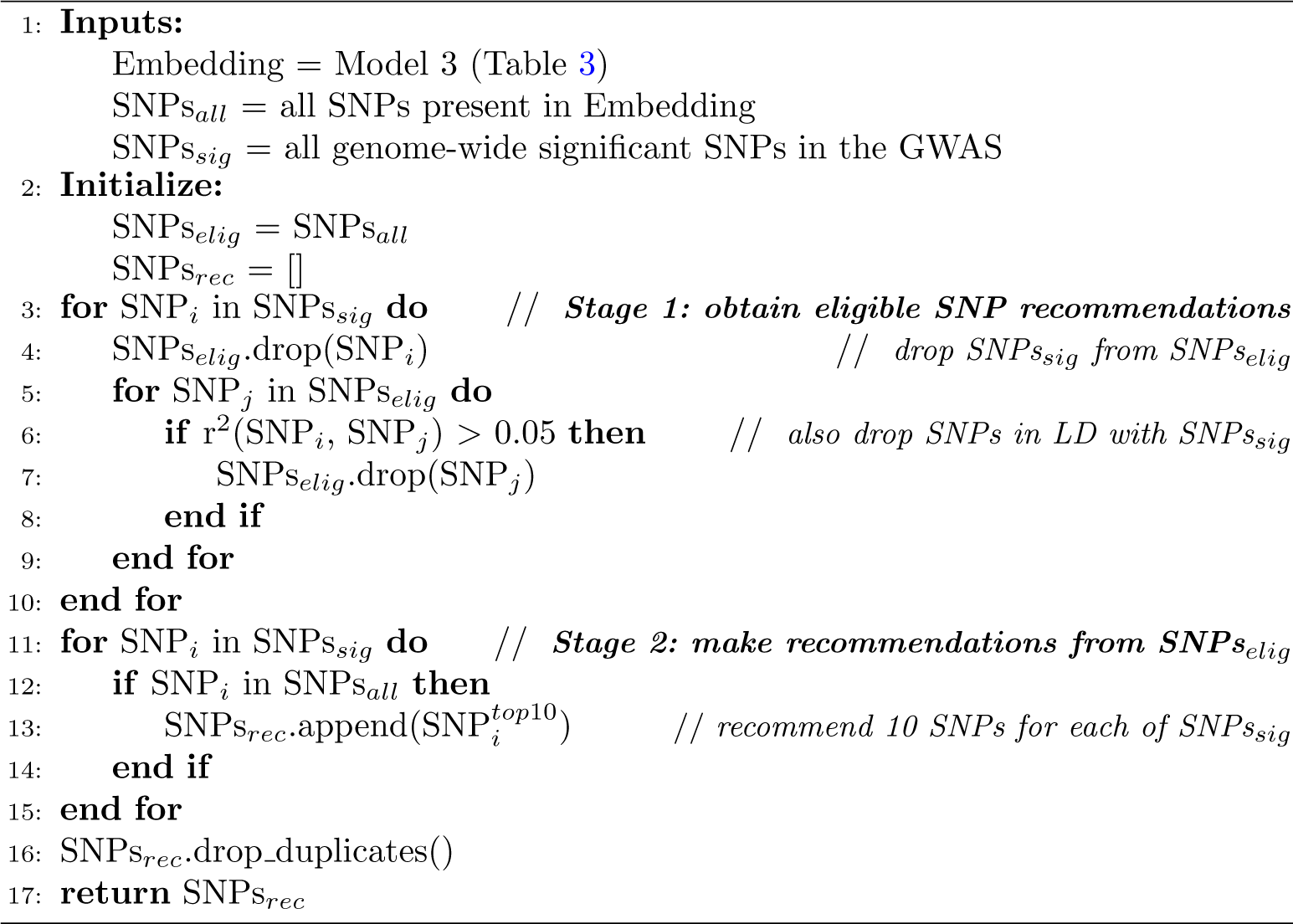

Following this, we performed an analogous analysis on the 64 diseases, identifying at least one novel locus in 64% (41/64). In total, we identify 143 independent novel disease associations. Excluding those involving the MHC region, this represents an uplift of 8% (128/1,548) compared to the number of independent associations that were identified at a genome-wide significant threshold in the GWAS performed in UK Biobank itself, only considering here those SNPs present in our embedding (Supplementary Table 3). In comparison, when an equivalent number of MAF-matched SNPs was randomly sampled per trait, only 25 independent associations across the 64 diseases reached statistical significance, demonstrating that we achieve an approximately six-fold improvement over random sampling. Figure 3 shows that the number of independent novel loci identified increases near linearly with the number of independent genome-wide significant associations identified in the original GWAS. For clarity, we refer in this manuscript to disease associations whereby the loci associated with each disease are considered independent (r^2^ < 0.05) as ‘independent associations’.

**Fig. 3.**
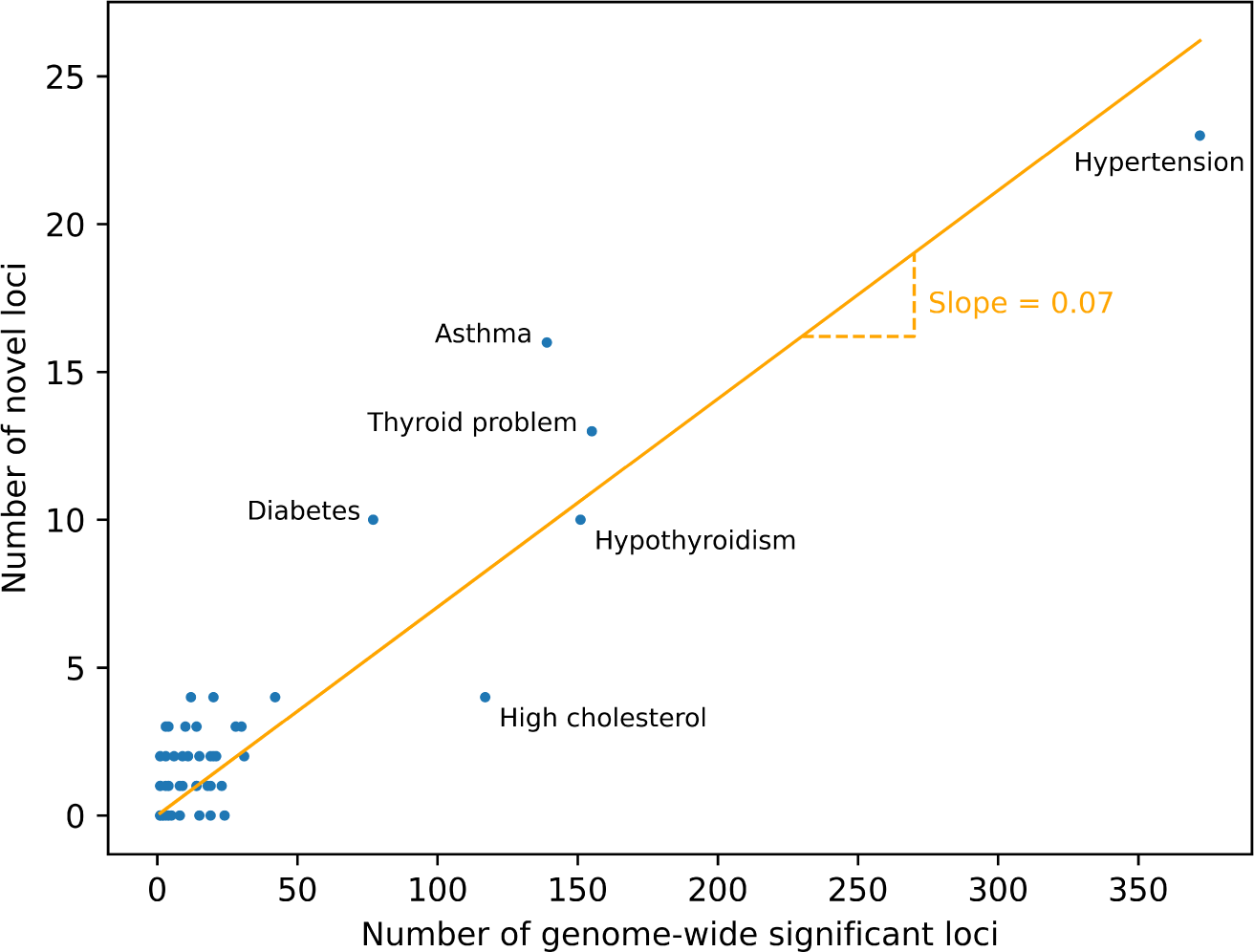
Complex traits are often extremely polygenic. The number of independent novel loci identified by our model increases approximately linearly (slope = 0.070, standard error = 0.003) with the number of independent genome-wide significant loci. 38% (39/103) of these novel loci were found to replicate in FinnGen (Main Text).

#### Supporting Evidence

Of the 143 independent novel disease associations, we attempted replication of 103 associations that were tested in FinnGen [7]. In the remaining cases (N = 40), the loci were either not reported in FinnGen, or were associated with disease outcomes that could not be mapped to a corresponding trait. Following per-trait Bonferroni correction, 39% (40/103) of the disease associations initially replicated in FinnGen, with 38% (39/103) also having a consistent direction-of-effect. Two of the replicated novel loci were located on a different chromosome from all genome-wide significant loci, six other loci were more than 1Mb from all genome-wide significant loci, and 14 were located between 100Kb and 1Mb away (Supplementary Table 3). Of those associations that were genome-wide significant in UK Biobank, 60% (729/1,212) replicated following per-trait Bonferroni correction. This larger discrepancy, in comparison to replication for height, is possibly explained by issues of statistical power and differing LD structure in the replication cohort.

There were four classes of replication quality: 1) where the novel association was genome-wide significant in FinnGen in its own right (p-value *<* 5 *×* 10^−8^; N = 10); 2) where the novel association passed a standard Bonferroni correction for the total number of tests for which replication was attempted, adjusting here for repeated replication attempts of nine novel diabetes-associated loci for Type 1 and Type 2 diabetes (p-value *<* 0.05*/*112; N = 33); 3) where the novel association passed a per trait Bonferroni correction (N = 39); and 4) where the result was not replicated at any of the above thresholds in FinnGen (N = 64).

Replicated associations that were genome-wide significant in their own right in FinnGen (class 1) included for instance that between rs10774625 (nearest gene: SH2B3/ATXN2) and coeliac disease (p = 1.3 × 10^−6^ in UK Biobank; p = 2.1 × 10^−10^ in FinnGen). An example of a replicated association that did not reach a genome-wide significant threshold in FinnGen (class 2 and class 3) was that between rs12350420 (nearest gene: MVB12B, for which rs12350420 is an eQTL [5]) and glaucoma (p = 1.0 × 10^−6^ in UK Biobank; p = 2.8 × 10^−7^ in FinnGen), for which orthogonal support exists from elsewhere [9].

We additionally examined those novel associations that did not replicate in FinnGen (class 4; N = 64). We searched the GWAS Catalog database [10] for evidence of genome-wide significance (p-value < 5 × 10^−8^). Acknowledging that the contribution of UK Biobank may not have been accounted for here, we mapped six additional loci to known disease associations for traits including asthma, hypertension and pulmonary embolism (Supplementary Table 3). Altogether, we found evidence in FinnGen or GWAS Catalog for 44% (45/103) of the novel disease associations we identified, none of which were genome-wide significant in UK Biobank.

As the ever-increasing study size of disease GWAS becomes accompanied by growing financial and logistical demands, we believe that our approach could play an important role in uncovering novel genetic disease associations, with the potential to enhance both past and future GWAS results.

## Methods

### Data

Molecular trait GWAS data: 1) meQTL: GoDMC [11]; and 2) eQTL: eQTLGen [5]. Non-molecular GWAS data: 1) height and diseases (discovery): GeneAtlas [12]; 2) height (replication): Yengo et al. [6]; 3) diseases (replication): FinnGen R9 [7]; 4) diseases (network centrality and supporting evidence): GWAS Catalog [10] (accessed 2 December 2022). LD reference cohort: EUR from 1,000 Genomes [13] (available here: https://mrcieu.github.io/ieugwasr/articles/localld.html; accessed 16 March 2023).

### LD clumping

We defined two sets of SNPs to consider based on the meQTL results in GoDMC: one pseudo-independent set and one dependent (standard) set. We first selected, from all the meQTL results in GoDMC [11], the maximum absolute z-score per genetic variant. We then performed LD clumping using Plink version 1.9 (https://www.cog-genomics.org/plink/) on the composite result. LD clumping was performed with the following settings: 1) r^2^ threshold of 0.05, and a flanking region of 1Mb around each index SNP (pseudo-independent set); and 2) r^2^ threshold of 0.9, and a flanking region of 1Mb around each index SNP (standard set). Due to the compute resource required, the standard set was limited to SNPs with a MAF > 0.05 (instead of a MAF > 0.01). 91,722 and 952,129 SNPs were retained in the pseudo-independent and standard sets, respectively. Analyses were based on the standard set, unless otherwise specified.

### Models

We used the Deep Graph Library (DGL; https://github.com/dmlc/dgl; version 1.0.0) implementation of the PinSage algorithm [4]. Models, together with their training and validation/test sets, are listed in Table 3.

**Table 3.** Model training and validation/test set.

|  | Training set | Validation/Test set |
| --- | --- | --- |
| Model 1 | 81% meQTL | 9% meQTL (validation); 10% meQTL (test) |
| Model 2 | 100% meQTL | 100% eQTL |
| Model 3 | 100% meQTL + 100% eQTL | non-molecular GWAS |

#### Model Training

Model training for the pseudo-independent set was performed on a NVIDIA TITAN X GPU using the following parameters: num-random-walks = 10, num-neighbors = 20, num-layers = 2, hidden-dims = 256, batch-size = 256, num-epochs = 300, batches-per-epoch = 5000 and lr = 5e-5. Model training for the standard set was performed on a NVIDIA A100 GPU using the following parameters: num-random-walks = 10, num-neighbors = 20, num-layers = 2, hidden-dims = 512, batch-size = 512, num-epochs = 200, batches-per-epoch = 5000 and lr = 5e-4. The generated embeddings are accordingly referred to as ‘pseudo-independent embedding’ and ‘standard embedding’.

#### Model Performance

The ‘hit-rate’ was measured in Models 1 and 2, and was obtained as follows. SNPs considered were limited to those present in the embedding. For each outcome trait (either CpG-site methylation or RNA expression) with more than one significantly associated (p-value < 5 × 10^−8^) SNP, the most significantly associated SNP, irrespective of whether a *cis* or *trans* QTL, was selected as the query SNP.

In order to generate a set of recommendations based on the query SNP, its similarity to all other SNPs in the embedding is computed and the resulting list ordered. Here, the similarity between two SNPs is given by the dot product of their respective embeddings – a more positive dot product between two SNP embeddings is indicative of greater similarity between the two SNPs. A given number of recommendations can then be made by truncating the ordered list at the desired point.

1, 10 or 100 recommendations were made using the model trained as per Table 3. If the recommendations contained at least one SNP that was also significantly (p-value < 5 × 10^−8^) associated with outcome trait, this was defined as a ‘hit’, and if not, a ‘miss’. The ‘hit-rate’ refers to the fraction of queried traits (CpG-sites or genes) for which at least one significantly associated SNP was identified. For Model 2, the ‘adjusted hit-rate’, whereby the same number of recommendations are made per RNA as there are SNPs associated to the RNA in the test set, is given by the proportion of SNP-RNA associations correctly predicted.

When predicting *trans* associations based on the lead *cis*-eQTL, genes were required to be associated with at least one *cis*-eQTL and at least one *trans*-eQTL. To compare model performance against chance, we permuted the labels and made recommendations based on these. We repeated this 1,000 times to calculate the mean number of hits obtained when making recommendations based on permuted labels. The reported p-value was based on a two-tailed Z-test according to the mean and standard deviation for the number of hits over the 1,000 trials.

### Network Centrality

Using the pseudo-independent embedding – given that estimation of eigenvector centrality would otherwise be computationally intractable – the similarity matrix was created by calculating the dot product of all pair-wise combinations of the SNP embeddings. The ‘importance’ of a given SNP based on the resulting matrix was measured using eigenvector centrality as implemented by the graph-tool library [14] (version 2.45). Since computation of the eigenvector centrality measure requires all entries in an adjacency matrix to be non-negative, absolute values of all matrix entries were taken as a pre-processing step, acknowledging the resulting matrix may not fully reflect relationships in the network.

We first filtered for genome-wide significant SNP-trait associations from GWAS Catalog, and retained only SNPs present in our pseudo-independent embedding. We then extracted a list of disease traits, which we assigned to several categories: autoimmune, cancer, cardiovascular, diabetes, neurological/psychiatric, obesity and a separate category (‘other’ diseases) for those that did not fit into any of these (Supplementary Table 1). We compared the eigenvector centrality of SNPs associated with these disease categories to those of approximately 10,000 randomly sampled SNPs from our embedding SNPs, with the same MAF proportions (the actual number of SNPs sampled deviated slightly from 10,000 in some cases due to numerical precision of the sampling implementation; range: 9,999 to 10,002).

### Novel Locus Identification

#### Height

##### Discovery – UK Biobank

We first removed from the set of eligible recommendations all SNPs associated with height at a genome-wide significant threshold (p-value < 5 × 10^−8^), as well as those in LD (r^2^ > 0.05) with them. Following this, we limited the SNPs to those present in our standard embedding. 10 recommendations were then made per genome-wide significant SNP. The recommendations were pooled and duplicates removed. The p-values of association with height in UK Biobank were then obtained from the original GWAS. The number of tests considered was defined as the number of unique SNPs in the set of recommendations. Accepting that it is a very conservative approach (given the lack of independence amongst recommended SNPs), these were only considered significant if their p-value passed a Bonferroni correction for this number of tests. We LD clumped (r^2^ threshold of 0.05, flanking region of 1Mb) those that were deemed significant in order to quantify the number of independent novel loci identified.

##### Replication – Yengo et al

In order to avoid issues with low statistical power in the replication cohort and assess the true replication rate of the novel associations we identified, we took advantage of a recently published ‘saturated’ map of the effects of common genetic variants for height [6]. Thus, we attempted replication of the independent novel associations identified (as per above approach) in the meta-analysis results of Yengo et al., where the contribution of UK Biobank was excluded by the original authors [6] (available here: https://cnsgenomics.com/data/giant2022/excludingUKB/; accessed 5 January 2024). We considered as significant those that passed a Bonferroni correction based on the number of tests for which replication was attempted.

#### Disease traits

##### Discovery – UK Biobank

64 self-reported disease traits were considered from UK Biobank. These were those that had at least 1,000 cases and at least one significant (p-value < 5 × 10^−8^) disease-associated non-MHC region [15] SNP with an imputation score greater than 0.9, present in the standard embedding.

Per disease trait, we performed an analysis analogous to that described for height (above), performing LD clumping to identify the number of independent novel loci. In order to quantify the uplift, we have excluded from the reported number of independent disease associations those linked to the MHC region (chromosome 6: 28,477,797-33,448,354) [15], given the challenge in estimating the number of independent loci in that region.

To compare our model performance against chance, we randomly sampled, per disease trait, MAF-matched SNPs equal to the number of recommendations made. The number of independent loci reaching statistical significance was obtained, as above, by applying a Bonferroni correction for the number of sampled SNPs, and LD clumping at a r^2^ threshold of 0.05. This procedure was repeated 100 times for each disease trait, and the mean number of independent loci reaching statistical significance calculated.

##### Replication – FinnGen

Replication of the independent novel loci was attempted in the FinnGen [7] cohort. UK Biobank disease traits were mapped to FinnGen in a one-to-one manner (except ‘diabetes’ which was matched to Type 1 and Type 2 diabetes), and excluded where no suitable trait was identified (Supplementary Table 3). FinnGen traits considered were those that had at least 1,000 cases. Loci were looked up in FinnGen via https://r9.finngen.fi/ (accessed 12 Aug 2023). Multiple-testing was considered either per trait or across all traits, and a Bonferroni correction performed accordingly.

## Supplementary Data

***Supplementary Figure S1:***

**Model 1 performance on meQTL test set over time.** Line graphs showing the improvement in Model 1 hit-rate over time when 1 (left), 10 (middle) or 100 (right) recommendations are made. Model performance is provided for both the standard (top) and pseudo-independent (bottom) embeddings. The hit-rate was evaluated every 50 epochs. The initial hit-rate (epoch 0) is based on initialised model weights.

***Supplementary Figure S2:***

**Similarity matrix heatmaps for all chromosomes.** Heatmap of the similarity matrix representing functional interaction across SNPs, for all 22 chromosomes. Genome position is on both axes. Higher values (darker red) indicate greater similarity between two loci, similarity here being given by the dot product of their respective embeddings. The diagonal represents self-similarity. High similarity scores closer to the diagonal represent the local (*cis*) region; whilst those further away from the diagonal represent *trans*-region(s) co-associating together. The similarity matrix is based on the pseudo-independent embedding.

***Supplementary Table 1:***

**Disease groupings for network centrality assessment.** List of traits belonging to each of the different disease categories considered when assessing network centrality.

**Supplementary Table 2:**
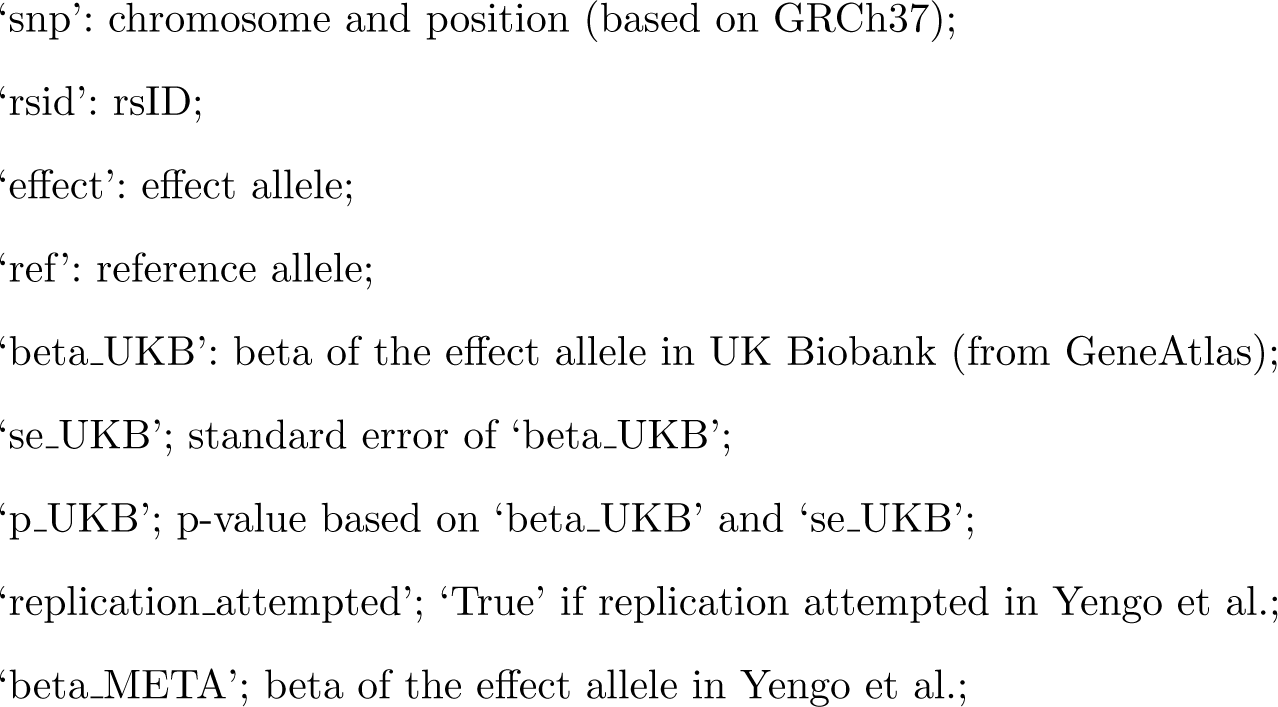
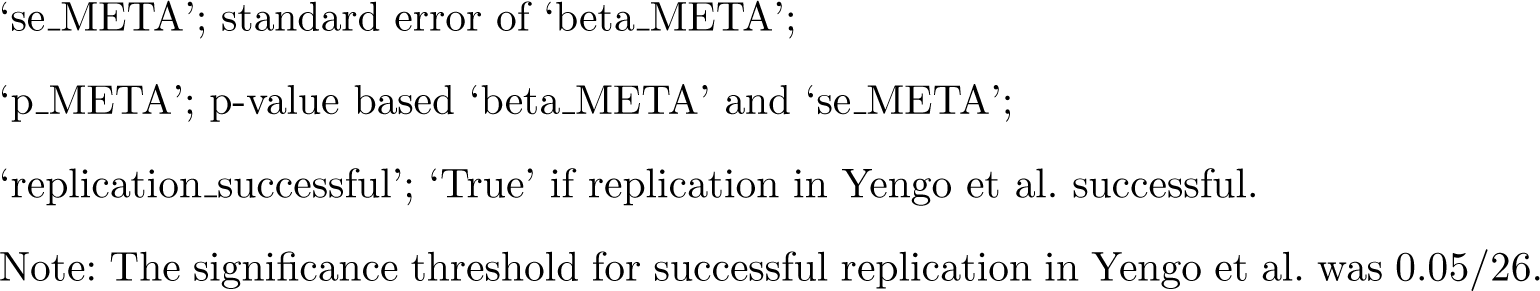
Novel height-associated loci and replication in Yengo et al. [6].

**Supplementary Table 3:**
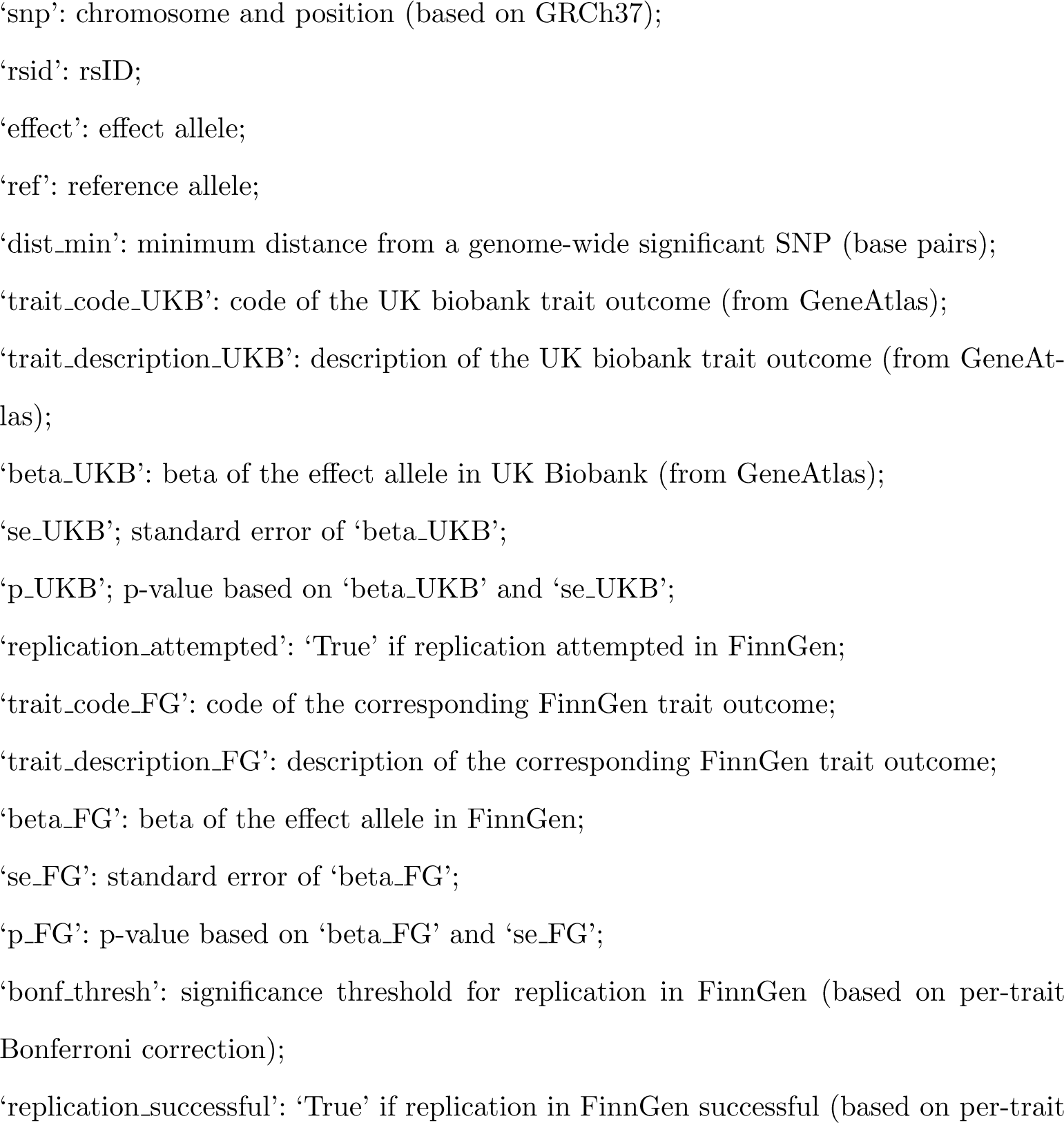
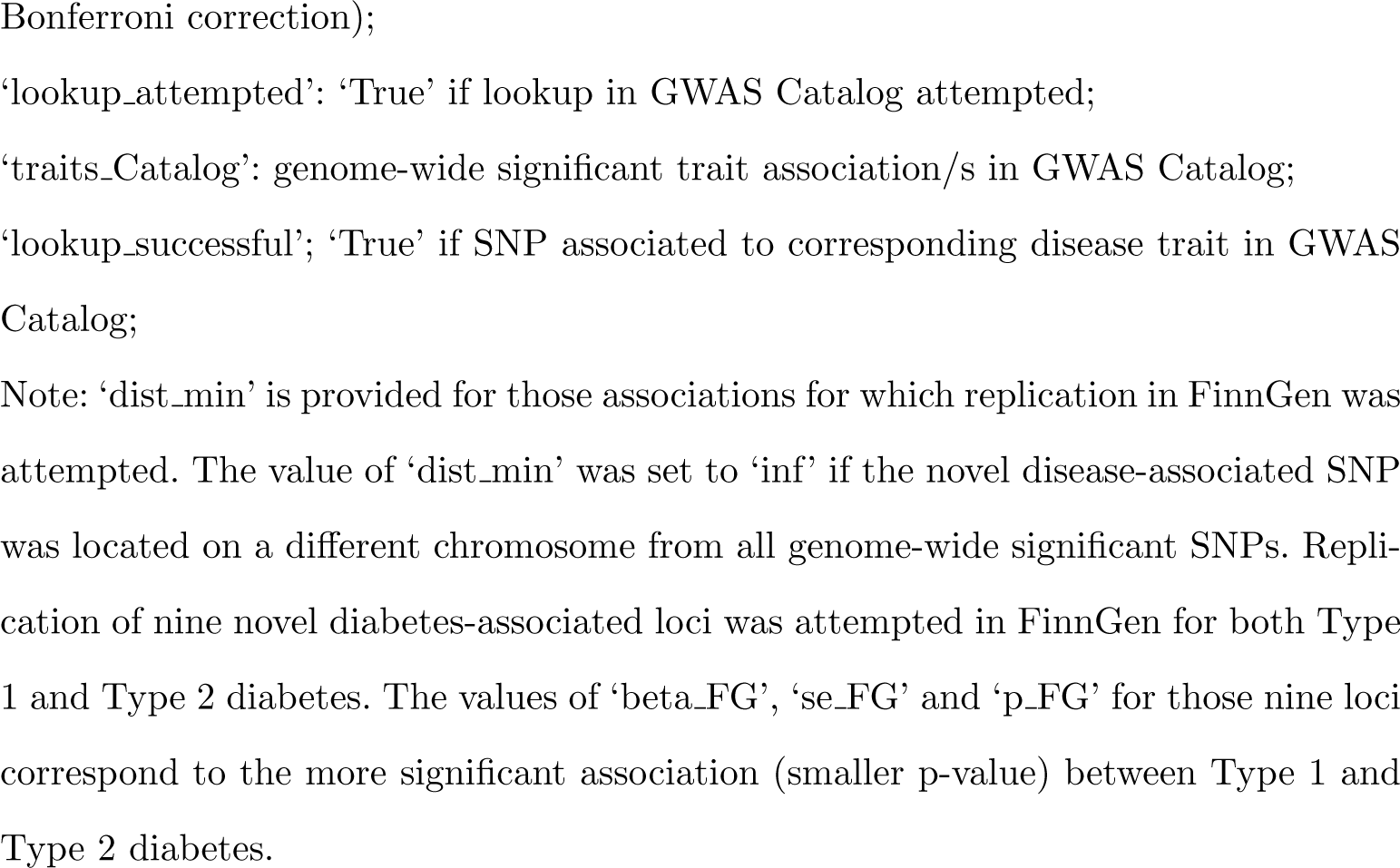
Novel disease-associated loci and replication in FinnGen and GWAS Catalog.

## Data Availability

All training/test and replication data used in this study are publicly available. The ‘standard’ and ‘pseudo-independent’ embeddings (meQTL only, and meQTL plus eQTL) – from which SNP similarity can be computed – are available at: https://doi.org/10.7488/ds/7689.

## Ethics

Ethical review and approval was provided by the Edinburgh Medical School Research Ethics Committee (REC Reference: 23-EMREC-052).

## Acknowledgements

We would like to acknowledge the participants and investigators of all the projects and studies from which we have used results: UK Biobank (GeneAtlas), the FinnGen study, GoDMC, eQTLGen, Yengo et al., the 1000 Genomes Project and GWAS Catalog.

