## Supplementary figures and images for "Leveraging molecular-QTL co-association to predict novel disease-associated genetic loci using a graph convolutional neural network"

### Supplementary Figure S1

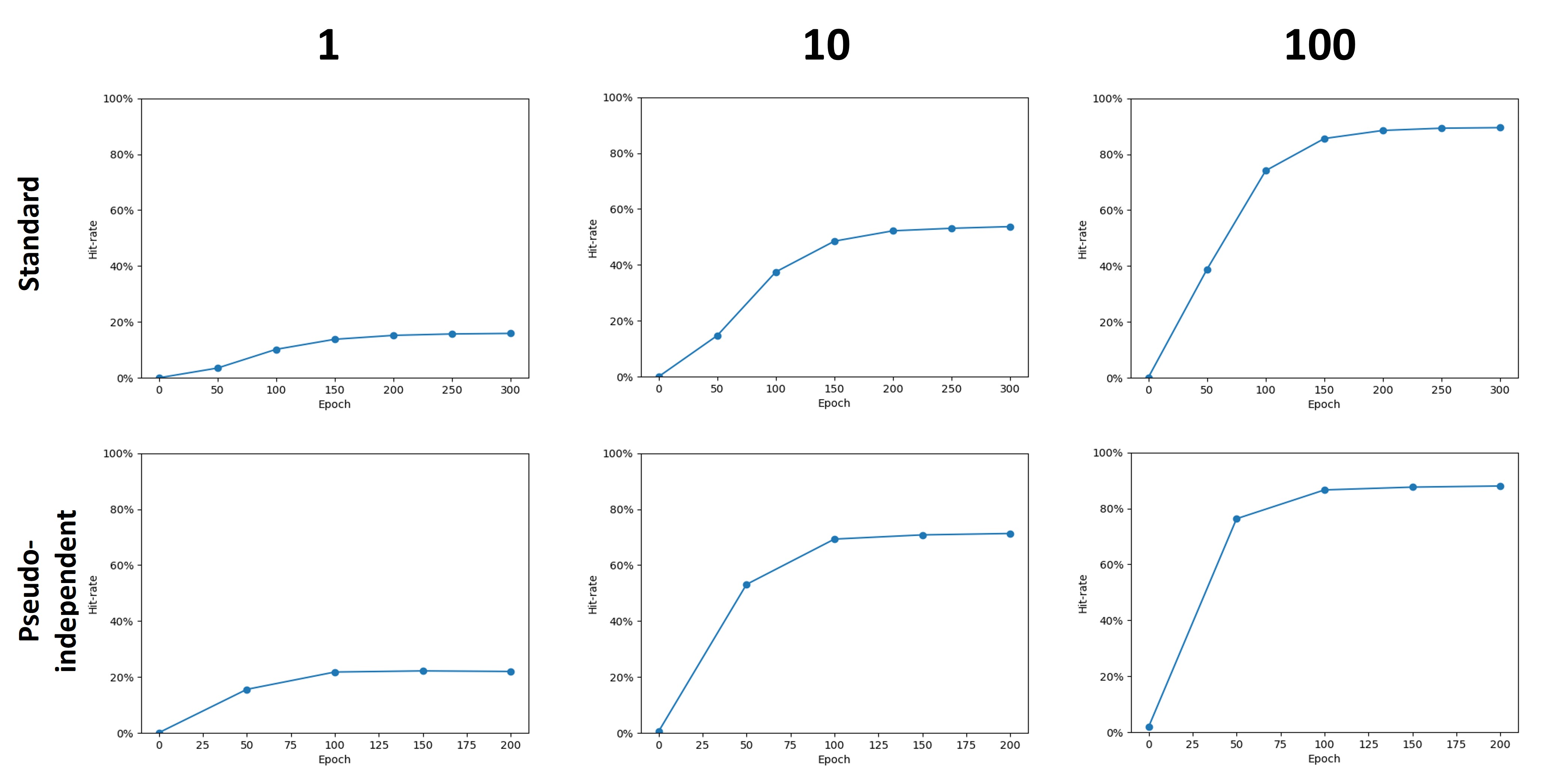

### Supplementary Figure S2

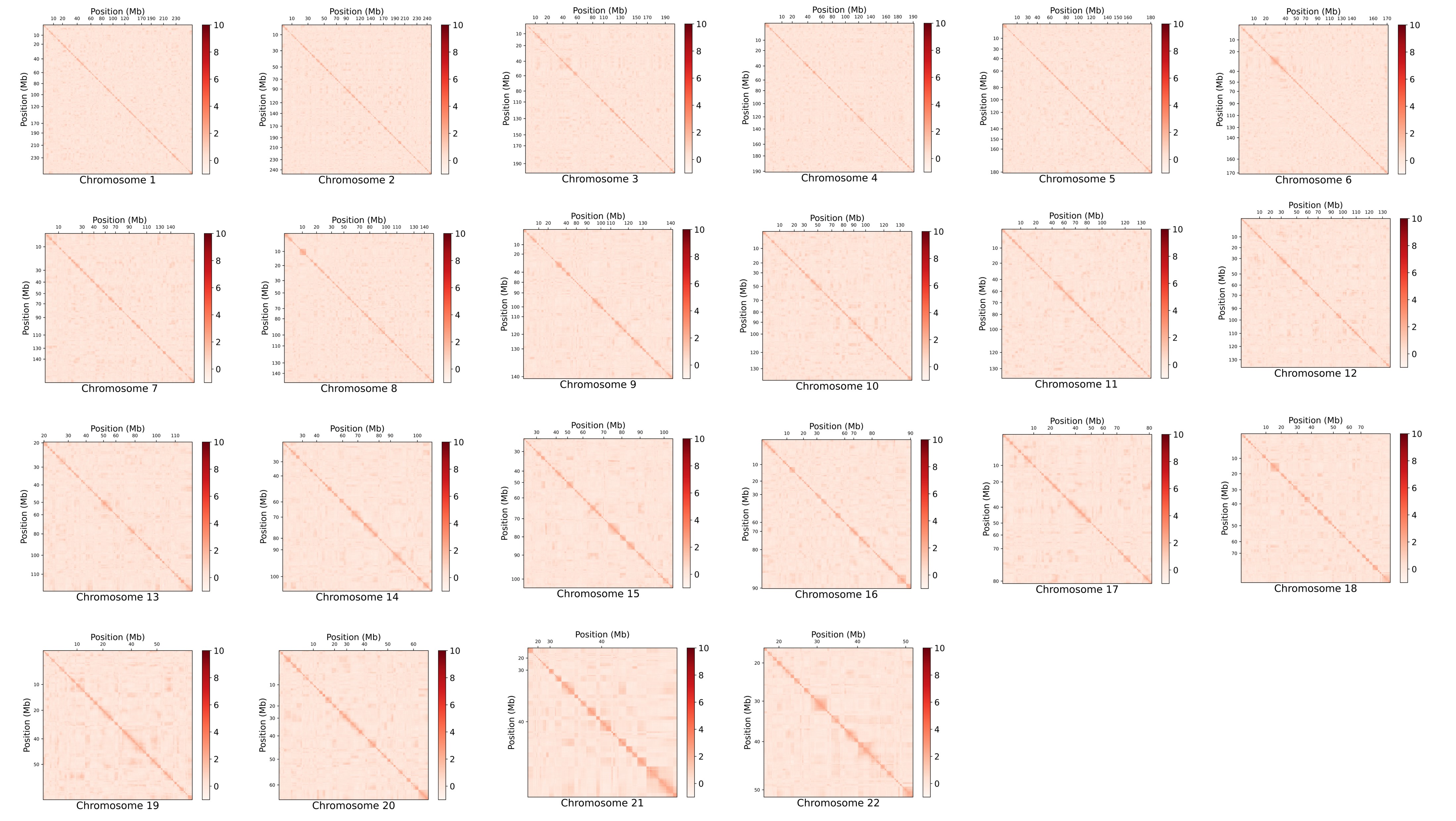
